# Factors influencing COVID-19 Infection in older individuals: History of Alcohol Use Disorder, Major Depressive illness, genetic variation and current use of alcohol

**DOI:** 10.1101/2021.12.06.21267386

**Authors:** Shirley Y. Hill, Brian J. Holmes, Jeannette Locke-Wellman

**Author notes:** Corresponding author: Shirley Y. Hill, PhD, Department of Psychiatry, 3811 O’Hara St., Pittsburgh, PA, 15213.

## Abstract

**Introduction:** The Coronavirus Disease 2019 (COVID-19) pandemic continues to be a major public health problem. Vulnerable populations include older individuals with presumed weakening of the immune response. Identification of factors influencing COVID-19 infection could provide an additional means for protecting such individuals.

**Methods:** Members of a family study previously interviewed as middle aged individuals were re-contacted and asked to participate in extended phone interview (2-3 hours) covering past and current mental health issues, physical health diagnoses, use of alcohol and drugs, and exposure to anyone with COVID-19. The average follow-up period was 32 years. Detailed medication use was collected to confirm medical diagnoses and to reveal possible protective effects of particular drug classes currently prescribed for the participant by their physician. Serology was available for red cell antigens (ABO, Kell, Duffy, Kidd, Rhesus) and HLA subtypes. Analyses were conducted to contrast COVID-19 + and COVID-19 - individuals for physical and mental health diagnoses, use of alcohol and drugs, and red cell and HLA serology. Additionally, analyses were conducted to contrast these groups with a group reporting known exposure but absence of COVID-19 symptoms or diagnosis by a health professional.

**Results:** Interviews were completed between September 2020 and November 2021. A total of 42 of the 90 individuals interviewed had been vaccinated at the time of interview. At the time of interview, 11.1% reported having developed COVID-19.

Using quantity per occasion (QPO) and quantity by frequency (QXF) totals in the past month by type of alcohol consumed, we found a significant association between QPO for liquor (p=0.017) and marginal effects for QXF for liquor consumption (p=0.06). Exposed individuals who were COVID-19 negative tended to drink more liquor than those who were positive, an average of about one drink per day. Beer and wine consumption were not statistically significant. A diagnosis of alcohol use disorder at baseline evaluation was not a significant predictor of being COVID positive or negative.

Self-reported current depression or depression in the past only was not a predictor of COVID-19 status based on a single question “Are you depressed currently or only in the past?”. In contrast, completion of a clinical interview designed to elicit depressed mood and concurrent symptoms for determination of the lifetime presence or absence of a depressive episode did reveal a significant effect. Comparison of responses at baseline to follow-up showed those most resilient to developing COVID-19 were those without evidence of a depressive episode by lifetime history at both points in time.

Physical health issues were analyzed for those that were frequently occurring in our sample such as hypertension but not found to be significant. BMI was analyzed and found to be statistically non-significant.

Analysis of HLA variation across the whole sample did not reveal a significant association but among males two variants, A1 and B8, did show significant variation associated with COVID-19+ and COVID-19-status. Analyses of the red cell antigens revealed one significant red cell effect; Kidd genotypic variation was associated with COVID-19 status.

**Interpretation:** We tentatively conclude that use of specific types of alcohol, namely liquor, is associated with reduced frequency of COVID-19. However, the amount is low, averaging about 1 drink per day. Enlarged samples are needed to confirm these results. The finding that past history of alcohol use disorder does not increase likelihood of developing COVID-19 is important. It should be noted that the 34 individuals diagnosed with AUD at baseline had survived an average of 32 years in order to participate in the current interview suggesting they may be especially resilient to adverse health conditions. The finding that a single question designed to elicit the presence or absence of depressed mood either currently or in the past was not a risk factor for COVID-19 in contrast to report of a clinically significant past history of a depressive episode based on more extensive examination using DSM criteria is important. Results for the KIDD blood group are novel and warrant further investigation.

---

While vaccines provide the primary source for limiting the spread of COVID-19, there is an urgent need to identify additional factors associated with progression to COVID-19. The World Health Organization reports significant rates of infection and death in the US (38,091,213 cases and 635,606 deaths as of September 2, 2021) and world-wide (218,205, 951 cases and 4,526,583 deaths). Identification of factors associated with development of symptoms, recovery, or progression and death is needed especially for vulnerable populations. Increased transmissibility associated with the emergence of new variants (1) along with reported reduced sensitivity to vaccine-induced antibodies (2) further underscores the importance of the study of these factors.

The role of alcohol in the immune response has been studied extensively, with a consensus that alcohol use has an impact on the immune response (3-6). Importantly, the level and direction of the response may be related to both beverage type and amount of alcohol consumed (7). Moderate drinkers in comparison to abstainers show reduced frequency of the common cold (8). Moderate alcohol use (1-2 drinks per day) in those intentionally exposed to five different respiratory viruses resulted in decreased incidence of infection (9). Congruently, consumption of wine shows an inverse relationship to incidence of the common cold (10); and moderate consumption of beer with increased production of T cell cytokines (7).

Demographic factors, health status, and immunogenetic factors are expected to mediate the effects of alcohol use or misuse on the likelihood of susceptibility to the virus, and the degree and course of illness progression. Among hospitalized COVID-19 cases, hypertension, diabetes and obesity are often noted (11), conditions frequently seen with heavy use of alcohol. Those with past or current alcohol use disorders (AUD) may have a greater risk for adverse consequences of SARS-CoV-2 exposure but the effects of remission are currently unknown.

The present report utilized a cohort of participants over the age of 55 for whom extensive clinical data was collected at two points in time, first in middle age and again at an older age. Clinical data includes extensive alcohol and other drug use histories, psychiatric diagnoses, and health histories. The second interview included reported exposure to individuals with SARS-CoV-2 or COVID-19 symptoms. The second interview also covered alcohol use, psychiatric symptoms, health status and SARS-CoV-2 exposure history providing potential factors, both distal and proximal, that may contribute to infection and progression. Identifying risk and protective factors in those over age 55 is an important public health issue because of the greater morbidity and mortality from COVID-19.

## Methods

### Participants

Individuals were included if they had previously been assessed as part of a family study that included family members from families with multiple cases of alcohol use disorder or were members of control families selected for absence of AUD. Individuals from the proband generation or their parents were included resulting in sample of individuals between the ages of 55 and 103 years at follow up. All were living in their own homes and not in a group setting such as a nursing home. To facilitate cooperation, all individuals were offered phone interviews for completion of the study.

### Clinical Assessment at Baseline

All participants signed University of Pittsburgh Institutional Review Board (IRB) approved Consent forms, following explanation of the study requirements and goals. All agreed to future contact. At baseline, psychiatric diagnoses and family history of alcohol dependence were obtained using a structured psychiatric interview (Diagnostic Interview Schedule [DIS]) (12) that provided Diagnostic and Statistical Manual of Mental Disorders III (DSM-III) diagnoses and quantity-frequency data from the Lifetime Drinking History, an instrument with good re-test reliability and validity (13). This data provided baseline diagnoses of alcohol use disorder for evaluating its presence as a pre-existing condition. Data from an extensive health questionnaire covering lifetime history of illnesses, accidents, and hospitalizations provided information on physical health status at baseline.

### Follow-up Interview

Following introductory letters and a positive indication of interest, all subjects were mailed a consent form approved by the University of Pittsburgh Institutional Review Board (IRB) outlining the material to be covered and requesting return of the signed consent form. At follow up, the same health questionnaire given at baseline was administered. The interviewer obtained dates of onset of health problems and whether the condition was currently present. Medication use with dose and duration of use was collected for verification of health status. All participants were queried about their frequency of alcohol use, usual and maximal quantity per occasion in the last 30 days and the past week. Specific type of beverage, beer, wine or liquor by frequency and quantity was also collected, requesting the informant provide “typical” and “maximal” quantity. An extensive history of current use of medications extending through the onset of the COVID was obtained. Those meeting a diagnosis of alcohol use disorder at baseline were interviewed with the portions of the DIS instrument to determine the current presence of problems associated with alcohol use.

#### Mental Health

### Depression

In the health and medication review, participants were queried about whether they were currently depressed or if only in the past using a single question. Later portions of the interview included detailed information concerning presence of low mood lasting 2 weeks or more accompanied by neurovegatative symptoms (e.g., loss of appetite) needed for presence or absence of a DSM-III diagnosis (the DSM criteria in place at baseline) of major depressive episode.

### Alcohol Use Disorder

Participants were asked to respond to questions allowing the determination of whether or not they met Feighner criteria (14). This procedure was followed so that comparison with the baseline interview which contained these questions and determination of whether the participant met the criteria could be compared across time periods. The Lifetime Drinking History questionnaire was also administered to quantify the amount of use in the most recent drinking epoch.

### Exposure and Illness

Respondents were asked to provide information on whether or not any person in their home had developed COVID-19 since the beginning of the pandemic (January 2020). Whether or not the individual experienced symptoms of COVID-19, and their level of involvement with the health care system was also determined.

### Immunogenetic Factors

Among the hypothesized mechanisms responsible for generating an immune response against Sars-Cov-2 include variation in the human leukocyte antigen (HLA) complex. These cell surface molecules provide an essential role in the recognition of non-self molecules by the acquired immune system. Antigens from the invading pathogens are bound by the HLA molecules facilitating the presentation of these to relevant T lymphocytes for initiation of an immune response (15, 16). HLA serology was available for all of the participants in this report.

### Red Blood Cell (RBC) Groups

Red blood cell antigens lie on the surface of RBCs. They are defined serologically by reagent sera that react with the antigen to produce blood cell agglutination. The data set available for analysis included the ABO, Duffy, Kell, Kidd, and Rh systems. Previous reports have suggested variations in ABO are associated with COVID-19 susceptibility (17,18). In one study (18) genome wide significance was seen for a locus on chromosome 9 near the ABO gene. Replication of these results have been found using large GWAS samples from the Ancestry DNA project data (19) and from BioBank samples (20). Associations for other red cell antigens have not been reported. Limited information is currently available for variation among individuals for non-ABO antigen systems and risk for COVID-19 though one study tested for non-ABO antibodies reporting a lack of association (21).

### Statistical Analysis

Based on the response to questions concerning known exposure to someone with COVID-19, a positive response to symptoms of COVID-19, or neither, individuals were classified into one of three groups: (1) Not exposed, (2) Exposed but COVID19 negative (E-COVID-19 -), and (3) Exposed and COVID positive (E-COVID-19+).

Because exposure is not consistently known, analyses were also completed using a two group dichotomy of COVID-19- (Groups 1 and 2 combined) versus COVID-19+ (Group 3) in addition. Analyses of quantitative data were analyzed by one-way analysis of variance. Binary variables were analyzed using a Chi-Square analysis and Fisher Exact analysis where appropriate. All data were examined for outliers. Analysis of known risk factors (e.g., BMI, hypertension) were analyzed using a 2 × 3 Chi Square analysis. Planned analyses were designed to include any variables from these analyses that showed statistically significant results in further analyses using binary log linear regression analysis to capture potentially important variation.

## Results

### Demographic Characteristics

Interviews were completed for 90 participants ranging in age from 55.6 to 103.3 years (Table 1). The sample included a greater proportion of females and those with higher socioeconomic status.

**Table 1.** Sample Characteristics – First 90 Cases.

|  |  |
| --- | --- |
| **Age at First Interview** |  |
| **Mean and Standard deviation** | **37.8 ± 7.9 years** |
| **Median** | **36.2 years** |
| **Range** | **23.1 - 70.0 years** |
| **Age at Second Interview** |  |
| **Mean and Standard deviation** | **68.5 ± 7.8 years** |
| **Median** | **67.7 years** |
| **Range** | **55.6 - 103.3 years** |
| **Education** | **14.7 ± 2.4 years** |
| **Socioeconomic Status (SES) <sup>a</sup>** | **42.7 ± 12.1** |
| **% Within top two levels** | **62.9%** |
| **% Within lower three levels** | **37.1%** |
| **Male** | **39** |
| **Female** | **51** |
| **Recruitment Risk Status - High Risk** | **61** |
| **Recruitment Risk Status - Low Risk** | **29** |
| **Vaccination Status at Interview - YES** | **42** |

<sup>a</sup> SES was determined using the Hollingshead method that combines occupational status with years of education and provides suggested numerical ranges for five SES levels. Hollingshead, A. A. (1975). Four-factor index of social status. Unpublished manuscript, Yale University, New Haven, CT.

### Alcohol Use

The present report focuses on recent use of alcohol (past month) due to better recollection of patterns of use for points closer in time to the interview. Data were also examined for lifetime use and use since the beginning of the pandemic (January 2020) to insure that recent use was reflective of the pattern of use that would have occurred during the pandemic. Results of the analysis (Table 2) show a significant association between quantity of liquor consumed in the past month and presence or absence of COVID-19. Similarly, the quantity per occasion differed by group for liquor consumption. Significant effects were not seen for beer or wine.

**Table 2.**
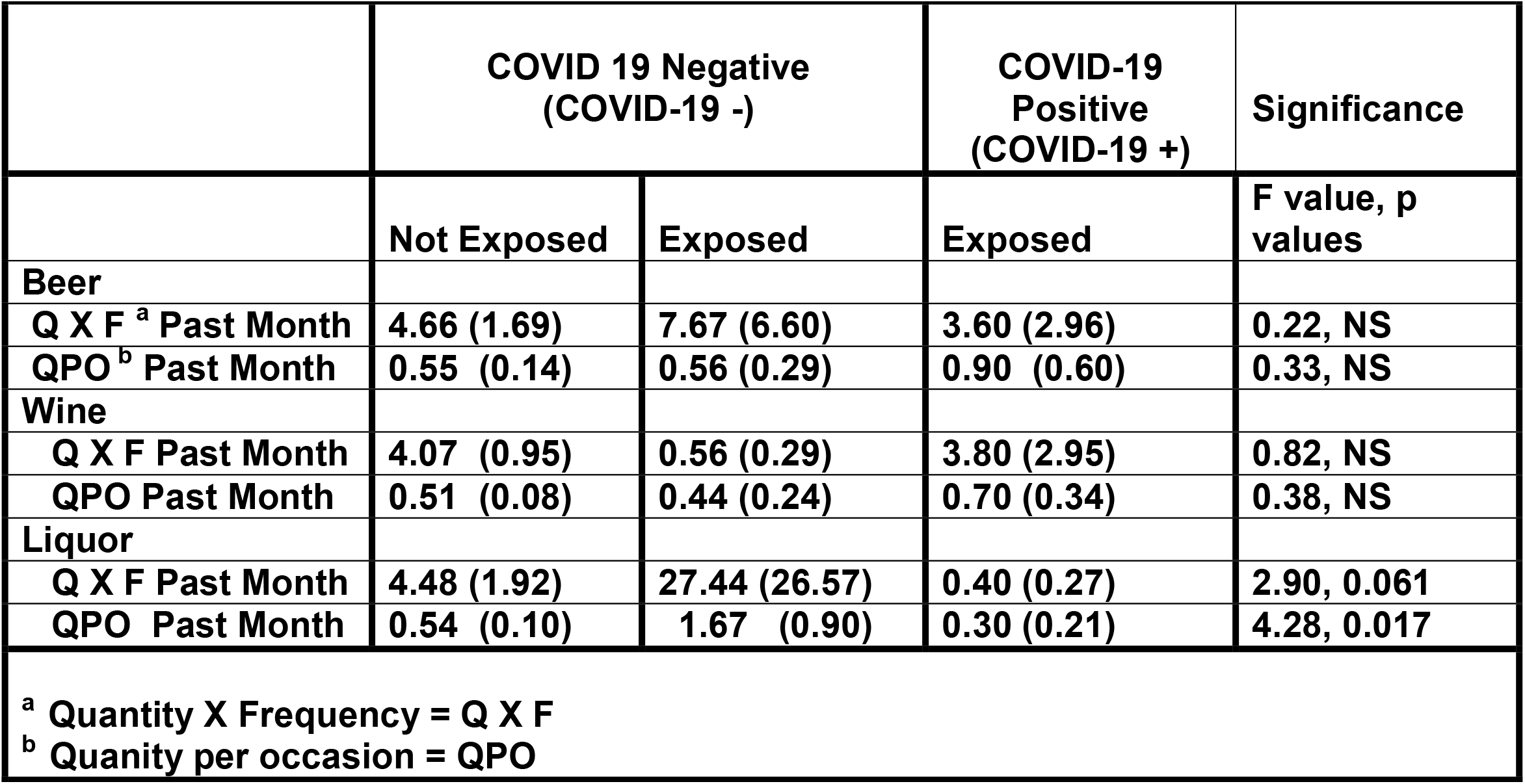
COVID-19: Mean and standard deviation for Not Exposed, Exposed-COVID Negative, Exposed-COVID Positive and Recent Alcohol Use.

### Mental Health

#### Alcohol Use Disorder

A total of 88 individuals were evaluated for presence or absence of a DSM-III diagnosis of alcohol dependence. Of these, 54 did not meet criteria and 34 met criteria at baseline. Previous diagnosis of an alcohol use disorder by DSM-III criteria or Feighner criteria at baseline was not associated with COVID-19 (Table 3). Similarly, meeting Feighner criteria for an alcohol use disorder at follow up was not associated with experiencing COVID-19.

**Table 3.** Alcohol Use Disorder (AUD) was determined with a DSM-III consensus diagnosis at baseline along with administration of questions needed to determine if Feighner Criteria were met at baseline. Feighner Criteria questions were given again at the 32 year follow-up.

|  | **DSM-III AUD PRESENT**<br>**AT BASELINE**<br>**(N=34)** |  | **DSM-III AUD NOT**<br>**PRESENT AT BASELINE**<br>**(N=54)** |  |
| --- | --- | --- | --- | --- |
|  | **COVID+** | **COVID -** | **COVID +** | **COVID -** |
| **DSM AUD PRESENT OR**<br>**ABSENT AT BASELINE**<br>**(N=88)<sup>a</sup>** | 4 (11.8%) | 30 (88.2%) | 6 (11.1%) | 48 (88.9%) |
| **FEIGHNER CRITERIA**<br>**EVALUATED AT BASELINE**<br>**(N=88)<sup>b,c</sup>** | 4 (12.1%)<sup>d</sup> | 29 (87.9%)<sup>d</sup> | 6 (11.3%)<sup>e</sup> | 45 (88.2%)<sup>e</sup> |
| **FEIGHNER CRITERIA**<br>**EVALUATED AT FOLLOW-UP**<br>**(N=67)<sup>f</sup>** | 4 (15.4%)<sup>d</sup> | 22 (84.6%)<sup>d</sup> | 4(12.9%)<sup>e</sup> | 27 (87.1%)<sup>e</sup> |

<sup>a</sup> Baseline presence of AUD was not associated with COVID-19.
<sup>b</sup> Feighner Criteria for AUD was not associated with COVID-19. All 88 participants with DSM III consensus diagnoses had Feighner Criteria information available.
<sup>c</sup> Cases with Feighner Criteria concordant with the DSM diagnosis are shown here (N=84). Note 3 cases seen as not having AUD did meet criteria for Feighner Criteria. Also, 1 case with AUD did not meet Feighner Criteria for AUD.
<sup>d</sup> Cases met Feighner criteria.
<sup>e</sup> Cases did not meet Feighner Criteria.
<sup>f</sup> 7 AUD cases at baseline did not meet Feighner Criteria at follow-up while 3 without AUD at baseline did meet Feighner criteria at follow-up.
<sup>g</sup> Statistical analysis was conducted first within each AUD group (present and absent) and then across the 4 groups. No statistically significant results were seen indicating lifetime diagnosis of AUD was not associated with presence or absence of COVID-19.

#### Depression

The **p**articipants’ responses to a single question concerning whether they were currently depressed, or if only in the past, elicited at the follow-up interview, was not significantly related to presence or absence of COVID-19 (Table 4a). In contrast, presence or absence of a lifetime occurrence of a depressive episode was significant (Table 4b).

**Table 4a.**
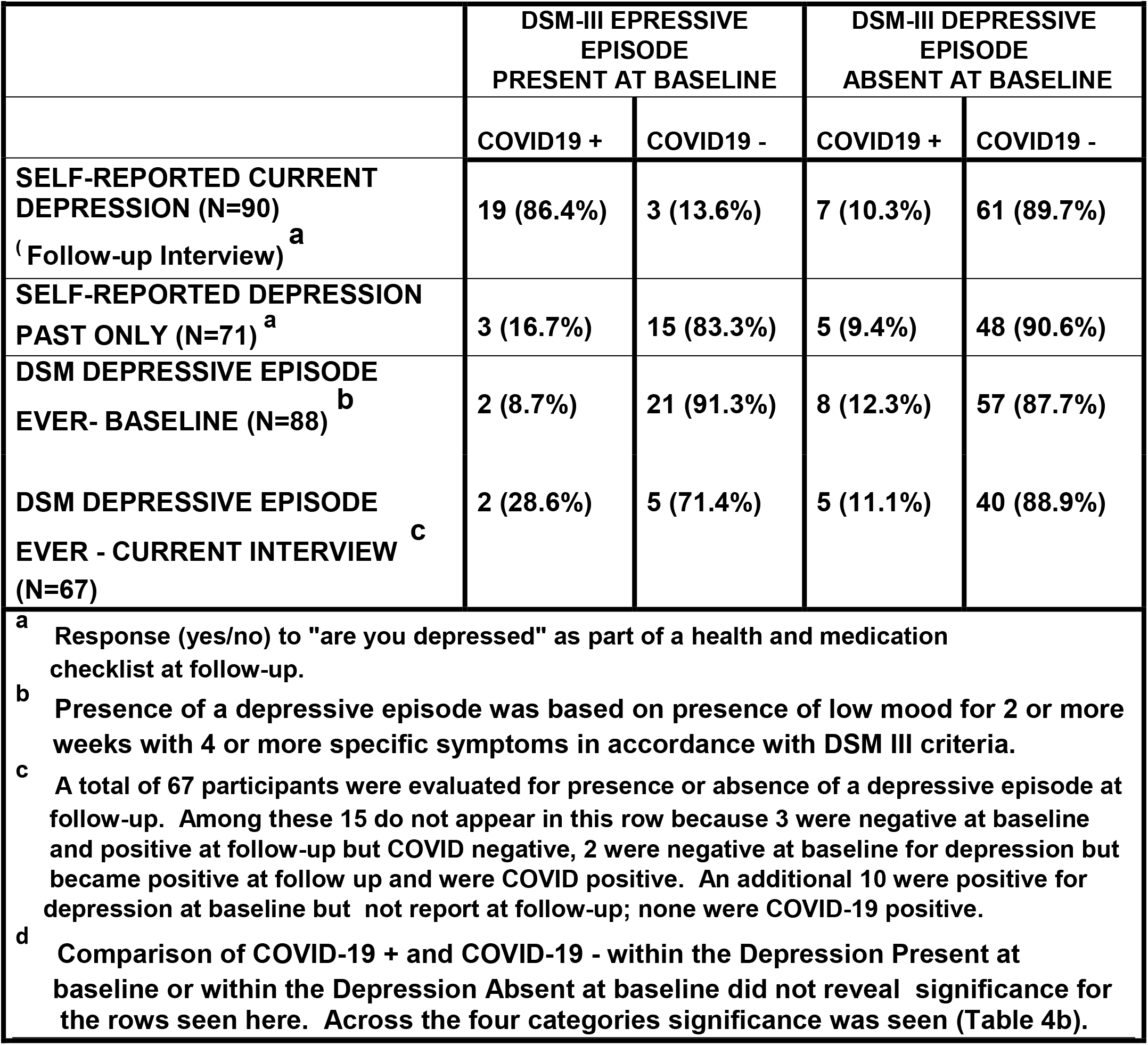
Self-Reported Depression ^a^ and DSM Diagnosis of Depressive Episode at Baseline and Follow-up.

**4b.** Presence of depressive episode reported at current interview and its association with COVID-19 presence or absence.

|  | COVID + | COVID - |
| --- | --- | --- |
| **Depressive Episode Present** | 4 (33.3%) | 8 (66.7%) |
| **Depressive Episode Absent** | 5 (9.1%) | 50 (90.9%) |

**Chi square value = 4.98, df=1, p=0.028; Fisher's Exact = 0.047.**

Determination of the presence or absence of a depressive episode was extracted from responses to the portion of the interview that included detailed information concerning presence of low mood lasting 2 weeks or more accompanied by neurovegatative symptoms (e.g., loss of appetite). Presence or absence of a DSM-III diagnosis of major depressive episode at baseline was determined in the same way.

### Stability and Change over 32 Years for Depressive Episodes

Presence or absence of COVID-19 in relation to a history of having a depressive episode was determined by contrasting three groups varying in the stability of the history depressive episodes. The influence of continued absence of a reported lifetime occurrence of a major depressive episode was evaluated by contrasting these participants (Group I) with those reporting a history of a depressive episode at follow-up independent of whether they reported an episode at baseline (Group 2) and those reporting an episode at baseline but now reporting no evidence of this (Group 3). A significant difference in the proportion of individuals with COVID-19 was seen in relation to the pattern of depressive episodes seen at two time points (Table 4c). Continued absence of a reported episode was associated with lesser likelihood of COVID-19 (Group 1). Current report of history of having a depressive episode resulted in a larger proportion with COVID-19 independent of baseline report (Group 2). Report of an episode at baseline followed by an absence at follow-up was associated with a reduced proportion with COVID-19 (Group 3)

**Table 4c.**
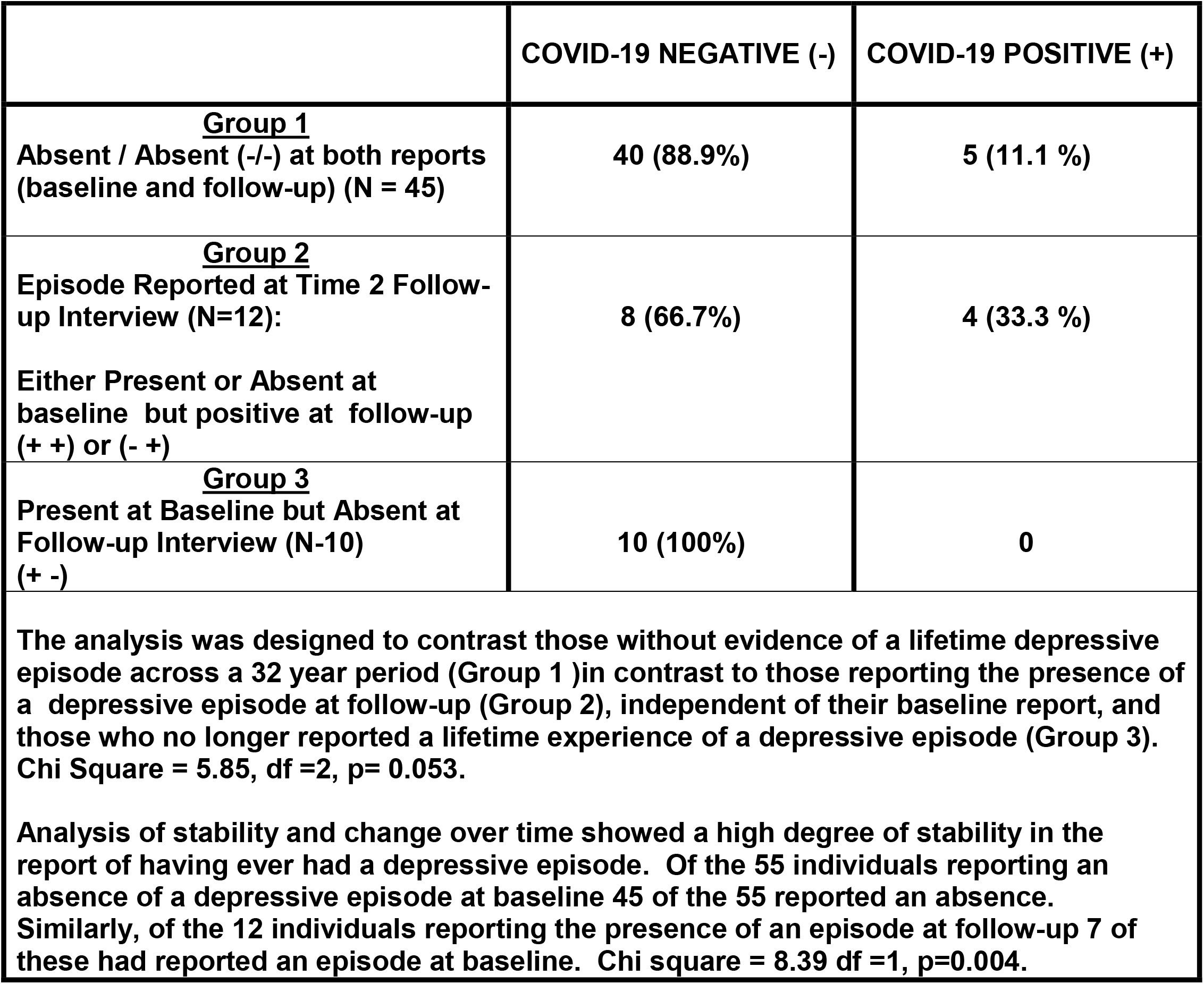
Depressive episode report across two interviews separated by 32 years.

### Physical Health

#### Hypertension

A number of studies have reported associations between hypertension and risk for developing COVID-19 (11, 22), with more severe consequences being reported in some studies (22). Based on reliable self report of hypertension in this sample (23), assessment of hypertension was completed using positive responses to whether or not the participant had been diagnosed with hypertension either currently or in the past. Using the combined report of either, a statistically significant association with reported COVID-19 was not seen. Among the COVID-19-group 63.7% reported past or current hypertension. For the COVID-19+ group 50% reported past or current hypertension with an equal number in each group.

Reports of both increased and decreased infection in those taking angiotensin converting inhibitor (ACE) medications for hypertension directed a further analysis of those reporting hypertension with use of ACE inhibitors. Overall, 18.9% of participants were currently taking an ACE inhibitor. Comparison of COVID-19+ and COVID-19-cases did not reveal a significant difference with 10% of those taking an ACE inhibitor reporting the presence of COVID-19 while 12.3 % of those not taking an ACE inhibitor reported having COVID-19 by the time of the follow-up interview.

### HLA and Red Cell Blood Types

#### HLA: Class 1 HLA variants were analyzed for all variants with sample frequency of 20% or greater

These included A1, A2, A3, B8, B44, and BW5. While no statistically significant associations were seen in the whole sample, two significant results were found in males (Table 5). Significant differences were found in males for A1 and B8. For A1, we find that among males the percentage with A1 who were COVID-19 negative was 23.5% whereas 80% of the COVID-19 positive carried A1 (Χ^2^ = 6.53, df =1, p=0.011). For B8, a trend was present for the whole sample (Χ^2^ = 2.77, df =1, p=0.096) with 25% of COVID-19 negative having B8 in contrast to 50% of the COVID-19 positive group. Among males only, the B8 results were significant (Χ^2^ = 8.89, df=1, p=0.003) with 17.6% of COVID-19 negative having B8 whereas 80% of the COVID-19 positive group carried B8.

**Table 5.**
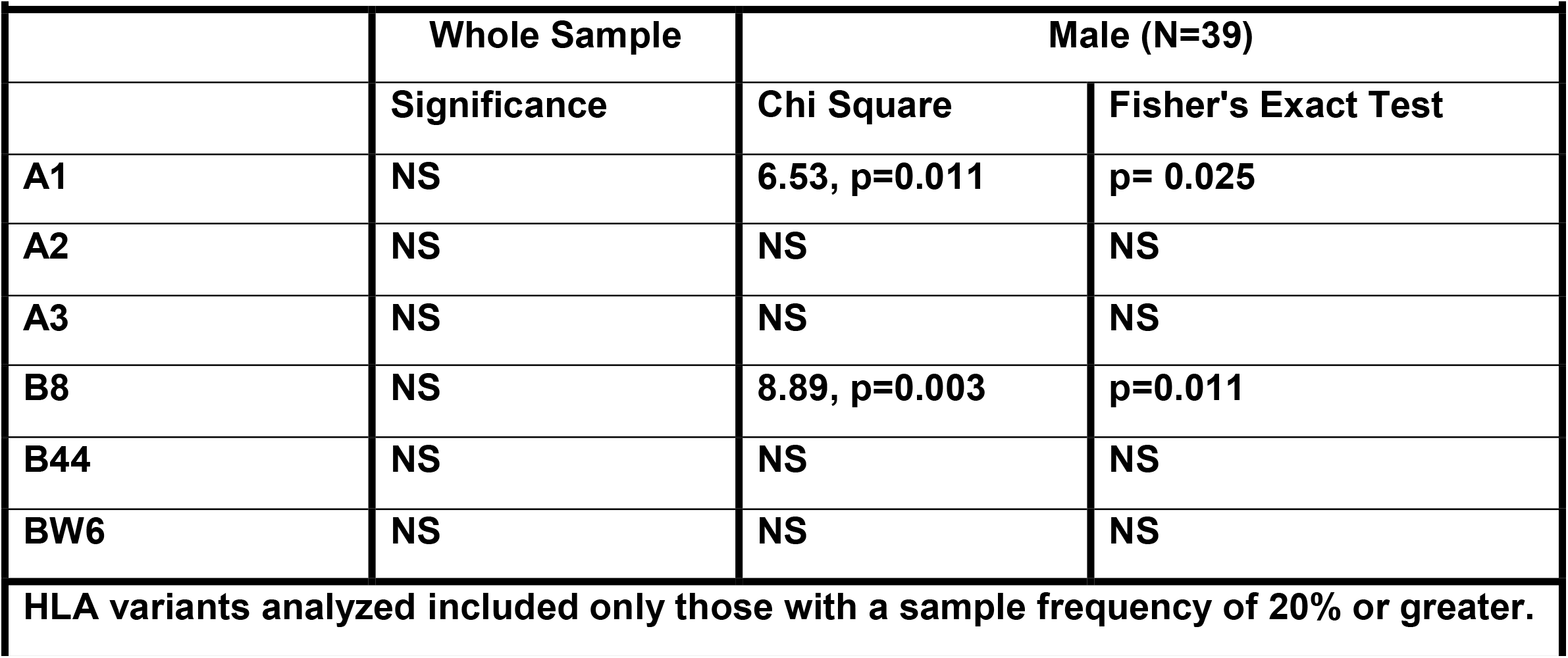
HLA Class I variants comparing COVID-19+ and COVID-19-cases.

|  | Whole Sample | Male (N=39) |  |
| --- | --- | --- | --- |
|  | Significance | Chi Square | Fisher's Exact Test |
| A1 | NS | 6.53, p=0.011 | p= 0.025 |
| A2 | NS | NS | NS |
| A3 | NS | NS | NS |
| B8 | NS | 8.89, p=0.003 | p=0.011 |
| B44 | NS | NS | NS |
| BW6 | NS | NS | NS |

HLA variants analyzed included only those with a sample frequency of 20% or greater.

#### Red Cell Blood Types

Comparison of COVID-19+ and COVID-19-groups for the Kidd red cell antigens revealed a significant difference by genotype Χ^2^ =7.38, df=2, p=0.025. Individuals who were homozygous for the non-dominant alleles appeared to have protection from developing COVID-19 (Table 6). Other red cell blood types were not associated with presence or absence of COVID-19 (Table 7). Additionally, the ABO group frequency among those reporting COVID-19 and those without did not differ significantly.

**Table 6.**
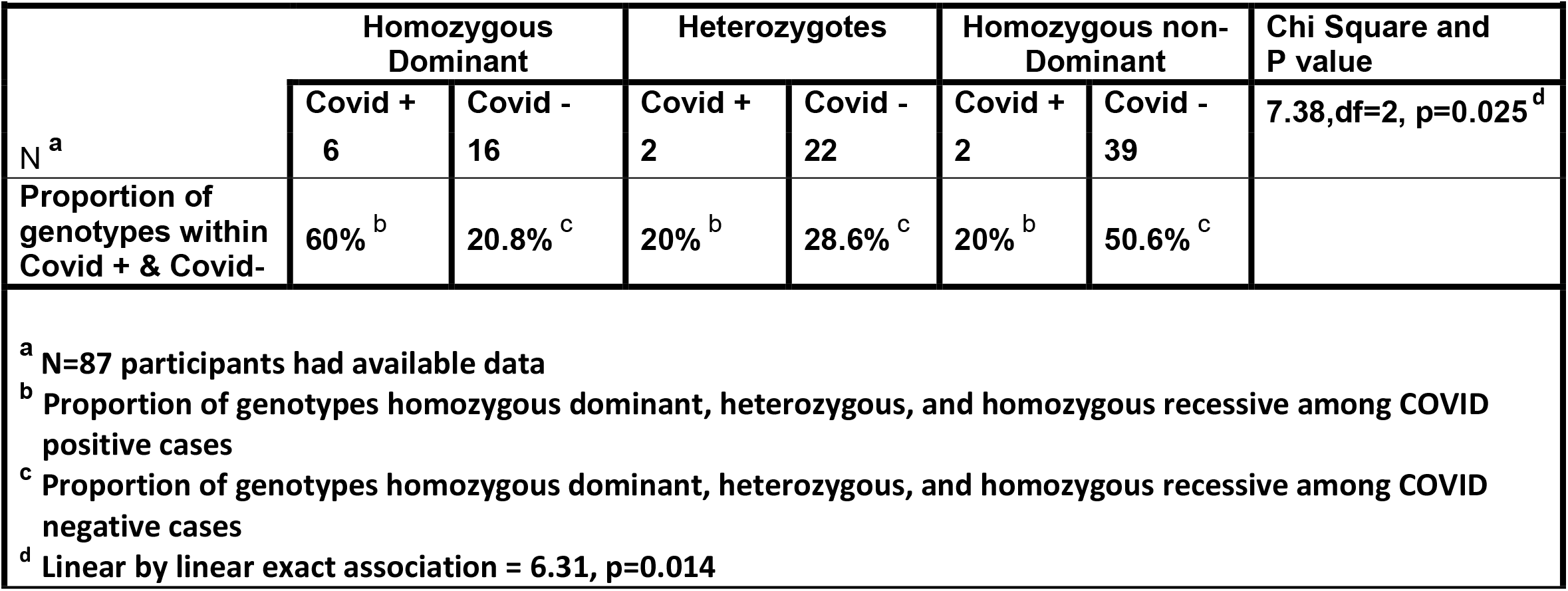
Kidd Red cell antigen and COVID-19 with suggestive evidence of a significant association.

**Table 7.** Red cell antigens with blood groups and presence or absence of COVID-19 at follow-up interview.

| RED CELL ANTIGEN | COVID NEGATIVE |  | COVID POSITIVE |  | Significance |
| --- | --- | --- | --- | --- | --- |
|  | PRESENT | ABSENT | PRESENT | ABSENT | Chi Square or Fisher's Exact |
| **ABO** |  |  |  |  |  |
| A | 28 (36.4%) | 49 (63.6%) | 4 (40%) | 6 (60%) | NS<sup>a</sup> |
| B | 14 (18.2%) | 63 (81.8%) | 1 (10%) | 9 (90%) | NS<sup>a</sup> |
| AB | 3 (3.9%) | 74 (98.1%) | 0 | 10 (100%) | NS<sup>a</sup> |
| O | 32 (41.6%) | 45 (58.4%) | 5 (50%) | 5 (50%) | NS<sup>a</sup> |
| **RH** |  |  |  |  |  |
| Positive | 59 (76.6%) | 18 (23.4%) | 8 (80.0%) | 2 (20%) | NS<sup>a</sup> |
| Negative | 18 (23.4%) | 59 (76.6%) | 2 (20.0%) | 8 (80.0%) |  |
| **Duffy** |  |  |  |  |  |
| Fy (a+b-) | 23 (30.3%) | 53 (69.7%) | 1 (10%) | 9 (90.0%) | NS<sup>b</sup> |
| Fy (a-b+) | 26 (34.2%) | 50 (65.8%) | 3 (30%) | 7 (70.0%) |  |
| Fy (a+b+) | 27 (35.5%) | 49 (64.5%) | 6 (60.0%) | 4 (40.0%) |  |
| **Kell** |  |  |  |  |  |
| KK | 1 (1.5%) | 65 (98.5%) | 0 | 8 (100%) | NS<sup>b</sup> |
| Kk | 8 (12.1%) | 58 (87.9%) | 0 | 8 (100%) |  |
| kk | 57 (86.4%) | 9 (13.6%) | 8 (100%) | 0 |  |

<sup>a</sup> Significance was tested using a 2 X 2 chi square analysis.
<sup>b</sup> Significance was tested using a 3 X 2 chi square (3 genotypes X 2 COVID conditions (positive or negative)).

## Discussion

The present report provides new information on the presence or absence of COVID-19 in association with pre-existing mental and physical health issues, current use of alcohol, and blood markers in individuals over the age of 55 during a 14 month period beginning approximately 9 months into the pandemic.

Among the strengths of the study is the longitudinal characterization of sample that enabled study of the impact of alcohol use disorder and presence of a major depressive disorder at baseline on outcome at a 32 year follow up. The results are encouraging in showing that individuals with a past history of problematic use of alcohol are not more vulnerable to developing COVID-19 based on this factor alone. Our results must be qualified by the fact that our participants as a group had relatively high socioeconomic status which may have provided overall better health care and outcome. The results also show the importance of careful evaluation of depressive disorders. No association was seen with COVID-19 outcome when those responding to a single question “Are you depressed?” were analyzed in contrast to finding that those with a major depressive episode having greater likelihood of experiencing COVID-19.

### Physical Health

Previous reports have noted that those with hypertension were at greater risk for developing COVID-19 and for experiencing greater severity if infected (11, 22). The absence of an association in the present study may be due to substantial use of antihypertensive medication in this sample suggesting the participants’ hypertension may have been well controlled.

### Alcohol Use

The present results appear to agree with previous studies (8, 9,10) showing improved immune functioning in those consuming low levels of alcohol consisting of approximately one drink per day (27.4 drinks in the past month). Although differences were found at the group level, we note significant individual variation in the quantity consumed. The type of alcohol consumed appears to be factor in any beneficial effect which alcohol may have on the immune system (7). The present results show specificity with associations being found for liquor but not for beer or wine. In considering an explanation for why differences might occur, it may be the case that consumption of liquor may lead to a faster rise in blood alcohol levels than beer or wine due to the need to consume larger liquid quantities over a longer period of time.

### HLA

The results found for B8 confirm previous reports that its presence was positively associated with incidence of SARS-CoV-2 infection (24). We found a trend for B8 to be associated with risk in the sample overall with this association becoming significant when evaluated in males only. Similarly, our results for A1 confirm previous reports of greater risk for infection if present (25). Our results differ from other reports in finding that A1 was a risk factor only in males.

### Mental Health and COVID-19

A previous report has found an association between mental health disorders and mortality among COVID-19 patients based on a meta-analysis of data from 7 countries (26). Among mental health disorders evaluated in one study, schizophrenia spectrum disorder was shown to be related to mortality among patients with COVID-19 though an association was not seen among patients with mood or anxiety disorders (27). Additionally, a large-scale meta-analysis did not find a relationship between preexisting mood disorders and risk for infection, hospitalization and death (28). In contrast, a recent analysis of COVID-19 infection and mortality found the odds of mortality was 2.78 times greater among patients testing positive who had a history of mood disorders (29). The study found this relationship independent of any medical comorbidities the patients may have had. Our results are in agreement with the study by Teixera et al (29) that found increased infection among those with a history of mood disorders.

Although previous studies have included a large number of participants, inconsistencies in results remain. All of the studies reported to date are based on cross-sectional analyses which may explain some of the variation. The present report though based on a small number of individuals has the advantage of data collection at two points in time. This has provided information on those who continue to report an absence of a mood disorders across a 32 year period and showing lesser rates of infection than those reporting a mood disorder at follow-up. The current analysis also allowed for determination of whether evaluation at the most recent point in time might be more salient in terms of risk for infection. Here, we find that those with no current report of a past episode at follow-up show proportionally fewer cases independent of a past report decades earlier.

Although there are considerable strengths in the methods used to uncover factors associated with developing COVID-19 in an older sample, we recognize there are limitations. These include the fact that we could not directly verify the presence of positive tests for SARS-CoV-2 infection but rather had to rely on the participants self-reports of testing positive and having symptoms and/or health care seeking for the symptoms. However, we believe that this cohort provides accurate responses to questions concerning health behaviors based on previous analyses in which the health behavior in question could be verified by in-person assessment of hypertension (23). An additional limitation was the form of HLA data available. Data had been collected using serological methods so that the allelic variation was not known. Nevertheless, we do find evidence for two variants as potential risk factors that are in accordance with other reports strengthening our conclusion that we were able to test HLA Class I variants for their phenotypic presence or absence.

In summary, the present report adds new information on the potential relationship between alcohol use during the pandemic, mental and physical health factors obtained over a 32 year span, along with genetic variation and SARS-CoV-2 infection. The sample studied included individuals over the age of 55 (55-103) residing in their own homes.

## Supporting information

STROBE Checklist

ICMJE Disclosure Form

## Data Availability

Per NIH guidelines data will be deposited at NIH.

## Acknowledgements

This is to express our sincere appreciation for an award from NIAAA/NIH for supporting this work (**3RO1 AA021746-05S1 Administrative Supplement to SYH)**. Also, we wish to thank the participants who patiently answered our questions in the phone interview that was conducted and for agreeing to participate in follow-up after a 32 year hiatus.

