## Supplementary material for "Factors influencing COVID-19 Infection in older individuals: History of Alcohol Use Disorder, Major Depressive illness, genetic variation and current use of alcohol": ICMJE Disclosure Form

| ICMJE DISCLOSURE FORM | |
| --- | --- |
| **Date:** | 12/5/2021 |
| **Your Name:** | Shirley Y. Hill, PhD |
| **Manuscript Title:** | Factors influencing COVID-19 infection in older individuals: History of Alcohol Use Disorder, Major Depressive illness, genetic variation and current use of alcohol3 |
| **Manuscript Number (if known):** | Click or tap here to enter text. |
| In the interest of transparency, we ask you to disclose all relationships/activities/interests listed below that are related to the content of your manuscript. “Related” means any relation with for-profit or not-for-profit third parties whose interests may be affected by the content of the manuscript. Disclosure represents a commitment to transparency and does not necessarily indicate a bias. If you are in doubt about whether to list a relationship/activity/interest, it is preferable that you do so.  The author’s relationships/activities/interests should be defined broadly. For example, if your manuscript pertains to the epidemiology of hypertension, you should declare all relationships with manufacturers of antihypertensive medication, even if that medication is not mentioned in the manuscript.  In item #1 below, report all support for the work reported in this manuscript without time limit. For all other items, the time frame for disclosure is the past 36 months. | |

|  | | | **Name all entities with whom you have this relationship or indicate none (add rows as needed)** | **Specifications/Comments (e.g., if payments were made to you or to your institution)** |
| --- | --- | --- | --- | --- |
| **Time frame: Since the initial planning of the work** | | | | |
| **1** | All support for the present manuscript (e.g., funding, provision of study materials, medical writing, article processing charges, etc.)  **No time limit for this item.** | | ☐ \| **None** 3R01 AA021746-05S1 \|; \| Click the tab key to add additional rows. | |
| **Time frame: past 36 months** | | | | |
| **2** | | Grants or contracts from any entity (if not indicated in item #1 above). | ☒ \| **None** | |
| **3** | | Royalties or licenses | ☒ \| **None** | |
| **4** | | Consulting fees | ☒ \| **None** | |
| **5** | | Payment or honoraria for lectures, presentations, speakers bureaus, manuscript writing or educational events | ☒ \| **None** | |
| **6** | | Payment for expert testimony | ☒ \| **None** | |
| **7** | | Support for attending meetings and/or travel | ☒ \| **None** | |
| **8** | | Patents planned, issued or pending | ☒ \| **None** | |
| **9** | | Participation on a Data Safety Monitoring Board or Advisory Board | ☒ \| **None** | |
| **10** | | Leadership or fiduciary role in other board, society, committee or advocacy group, paid or unpaid | ☒ \| **None** | |
| **11** | | Stock or stock options | ☒ \| **None** | |
| **12** | | Receipt of equipment, materials, drugs, medical writing, gifts or other services | ☒ \| **None** | |
| **13** | | Other financial or non-financial interests | ☒ \| **None** | |
| **Please place an “X” next to the following statement to indicate your agreement:** | | | | |
| ☒ | | I certify that I have answered every question and have not altered the wording of any of the questions on this form. | | |
